# Exploring Auditory Biofeedback Paradigms for Gait Training in Children with Cerebral Palsy: A User-Centered Design Study

**DOI:** 10.64898/2026.05.29.26353852

**Authors:** Prithvi Ravi Kantan, Mikkel B. Hansen, Jacob J. Foldager, Frederik S. Fjeldgaard, Sofia Dahl, Erika G. Spaich

## Abstract

**Purpose:** To identify, through iterative user-centered design, the auditory biofeedback requirements and sound preferences supporting gait training in children with cerebral palsy (CP), and to determine which feedback variables, sound mappings, and sound types yield clinically viable and movement-interpretable paradigms.

**Methods:** The iterative process spanned two prototype phases. Prototype A comprised seven paradigms demonstrated to two experienced physiotherapists (Workshop 1A). Two of these were subsequently discarded owing to poor sound-movement interpretability and two were modified. Six paradigms were added to Prototype B, demonstrated to four children, five parents, and one therapist (Workshop 1B) and two therapists (Workshop 2B). Data were analyzed using systematic text condensation.

**Results:** Within-child sound preferences varied with energy level and sensory state on a given day. Sound-movement interpretability tended to suffer for paradigms with greater acoustic complexity (e.g. computer-generated music). Therapists endorsed a repertoire spanning both movement quality and movement quantity targets. Participants independently proposed paradigms rewarding restrained and controlled movement, a feedback category absent from the current prototype.

**Conclusions:** Session-level calibration is preferable to fixed sound profiles, requiring real-time interface support for paradigm adjustment. Acoustic complexity must remain subordinate to movement-sound interpretability. Paradigms targeting movement restraint are a development priority unaddressed in the literature.

## 1 Introduction

*Cerebral palsy (CP)* describes a group of permanent disorders affecting movement and posture development, attributed to non-progressive disturbances in the developing fetal or infant brain [1]. As the most prevalent cause of childhood physical disability, CP affects approximately one in 500 newborns worldwide [2, 3]. The condition imposes substantial individual, familial, and societal burdens. For affected individuals, mobility is compromised due to motor and sensorimotor disturbances, with gait function representing a primary focus of therapeutic intervention to increase independence and mitigate long-term secondary complications spanning musculoskeletal, sensory, and cognitive domains [1, 2, 4].

The pathophysiology of gait impairment in CP is substantially attributable to disrupted sensorimotor integration across multiple levels of the neuraxis [5]. Typical locomotion demands continuous delivery of somatosensory signals from peripheral receptors to numerous supraspinal networks, which must coordinate motor output in real time to meet the variable demands of the environment [6]. Muscle and joint afferents from the leg, and tactile-kinesthetic afferents from the plantar surface are particularly central to this process: the afferent signals first engage spinal reflex circuits at the spinal level before ascending to the primary somatosensory cortex, where lower-limb afferent input is organized into a spatially structured representation of the body relative to the supporting surface [7–9]. The brain combines this feedback with feedforward signals derived from prior experience and visual, vestibular, and auditory information to predict and precisely plan subsequent steps [10–12]. In children with CP, the non-progressive brain lesion can disrupt the cortical and subcortical networks responsible for this integration, impairing their capacity to filter and weight sensory inputs, update internal body models, and generate the feedforward adjustments on which efficient gait depends [5]. Aberrant cortical oscillations are also coupled to secondary microstructural changes within the spinal cord, indicating that cortical disruption propagates downstream to impair the spinal interneuronal circuits on which coordinated locomotor output depends [13, 14]. Approximately 80 percent of children with CP experience sensory processing difficulties that manifest as decreased kinesthetic sensory processing and impaired visual perception [15, 16]. When combined with motor strategies such as increased and inappropriate co-activation of agonist and antagonist muscles [17], this creates a dual therapeutic challenge. The necessary learning process is demanding and requires clear, repeated sensory feedback, which is difficult when the child’s inherent capacity to integrate natural feedback may be compromised [18–20].

Physiotherapy constitutes the primary treatment modality and demonstrates documented efficacy in improving gait function among children with CP [21]. Treatment is grounded in motor learning principles, where repetition and intensity are critical for acquiring more appropriate movement strategies [21, 22]. However, the high repetition requirements can prove physically and mentally taxing, often reducing motivation and engagement [23]. As a means to address this, research suggests that children are generally more motivated by interactive and technological solutions [2, 24]. Technology-based approaches including biofeedback have been developed as supplements to traditional physiotherapy [25], and are capable of supporting motor learning by augmenting or supplementing sensory feedback [26]. Augmented feedback can help compensate for the sensorimotor limitations arising from cerebral injury, and has demonstrated effectiveness in supporting movement pattern correction and improving gait parameters in children with CP across several studies [27–30]. Recent reviews, however, indicate that technology-based gait rehabilitation for CP remains at a relatively nascent evidence stage. Both Mac-Intosh et al. (2019) and Schoenmaker et al. (2023) conclude that there is a need for further work comparing knowledge-of-performance and knowledge-of-results paradigms across both qualitative and quantitative methodologies [20, 31]. The most recent systematic review on augmented feedback for gait rehabilitation in CP, spanning 25 studies and 409 participants, identified consistent support only for velocity improvements and ankle kinematics, while evidence for step length, stride length, and cadence remained inconclusive due to methodological heterogeneity [30]. Despite these issues, the specific technique of auditory biofeedback has attracted particular attention within the field, driven by practical properties that suit the specific demands of gait training in children with CP.

As auditory biofeedback utilizes the medium of sound to communicate movement information, it can provide clear and immediate feedback without burdening the child’s visual attention [32], and requires minimal setup compared to systems relying on dedicated visual displays or haptic actuators [26]. This practical profile is further suited to the CP population by the substantial prevalence of cerebral visual impairment, estimated at 35–85% across the population, which limits the applicability of visual feedback for a considerable proportion of affected individuals [30]. Baram and Lenger demonstrated that a simple auditory solution using inertial sensors placed on the lower extremities, converting each step into a click sound delivered through headphones, achieved effects on gait-specific parameters comparable to visual biofeedback [33]. The design of the auditory signal itself nonetheless matters: evidence suggests that simple click sounds are insufficiently adapted to children’s motivational and sensory needs [24], and that more refined approaches yield better outcomes [31, 34]. Hamed et al. (2011) demonstrated that feedback designs incorporating rhythmic modulation of melodies can optimize effects on gait patterns [34], while Schoenmaker et al. (2023) found music-based auditory feedback to be associated with considerably larger effect sizes than click-based designs, proposing that richer stimulation may carry additional motivational value for children [31]. The authors nonetheless caution that the heterogeneity of study designs and outcome measures across the available literature precludes firm conclusions about the relative effectiveness of specific feedback forms [31].

Integrating technology into clinical gait rehabilitation for children with CP requires careful attention to how solutions are developed and adopted over time. Technology-based approaches have proven consistently engaging for children with CP [35], though sustained long-term use is closely tied to whether solutions are sufficiently calibrated, adaptable, and personally meaningful to the individual child [36, 37]. User-centered and iterative design, developed in close collaboration with children, families, and professionals, is the approach that best aligns technology with these requirements [24, 38, 39]. Technology development for this population is further complicated by substantial heterogeneity within the CP group: needs and preferences vary markedly even among children of similar age [40], and the relevant variation spans motor functional impairments as well as sensory, cognitive, and psychosocial factors that shape both engagement with technology and intervention outcomes [1, 41]. Children’s developmental profiles are also in constant flux, requiring flexible and adaptive solutions capable of accommodating changing needs, making meaningful involvement of children, parents, and professionals throughout the design and evaluation process essential rather than optional [39]. Children with CP bring specific contextual challenges to biofeedback use, including reduced attention span, motivational fluctuations, and sensory integration difficulties, each of which can affect how feedback is perceived and whether engagement holds across sessions [24, 42].

To ensure both effectiveness and usability, it is therefore critical that technologies for auditory feedback be developed in close collaboration with the children themselves, their families, and the professionals who will use the technology in practice [39, 41]. By systematically exploring the target group’s requirements and specifications, development can be tailored to the needs and therapeutic goals of the children, reducing the risk of research waste resulting from inadequate or misinformed design when overall effectiveness is to be documented [43]. An intervention design experienced as motivating and personally meaningful has been identified as a key prerequisite for sustaining the training engagement necessary to drive functional improvements in children with CP, though the direct evidence base linking specific motivational intervention features to measurable gait outcomes remains limited [44]. This gave rise to the present focus on investigating how children with CP experience sound-based biofeedback, and how this technology can be designed to support motor learning in a meaningful and motivating manner, grounded in existing evidence and established theoretical concepts [20, 27]. We aimed to address the following research questions:

1. What are the needs and preferences of children with CP, their families, and therapists with regard to auditory biofeedback technology for improving gait function?
2. What feedback variables, sound mapping structures, and sound types yield clinically viable and movement-interpretable auditory biofeedback for gait training in children with CP, as assessed by experienced physiotherapists?

## 2 Methods

### 2.1 Study Design

Building on the methodology applied in previous work [45, 46], the present study adopted an iterative framework in which workshops served as the primary mechanism for eliciting user input and findings from each workshop were systematically translated into design revisions before the subsequent evaluation round. Two workshop types were conducted across two prototype iterations (A and B), as depicted in Figure 1. Workshop 1 (sound preferences and needs) was conducted with children with CP, their parents, and therapists to explore auditory sound preferences within a gait training context, and to elicit feedback on which acoustic properties, sound types, and feedback behaviors participants found engaging, motivating, or appropriate for children with CP; this workshop type was designed to address RQ1. Workshop 2 (feedback paradigm evaluation) was conducted with experienced therapists to evaluate specific feedback paradigms developed on the basis of Workshop 1, assessing clinical utility, movement-sound intuitiveness, and suitability for therapeutic application in children with CP; this workshop type was designed to address RQ2. In the remainder of the paper, we use a labeling convention in which the numeral denotes the workshop type and the letter denotes the prototype version used, such that Workshop 1A refers to the Workshop 1 session conducted with Prototype A, and so forth.

**Figure 1:**
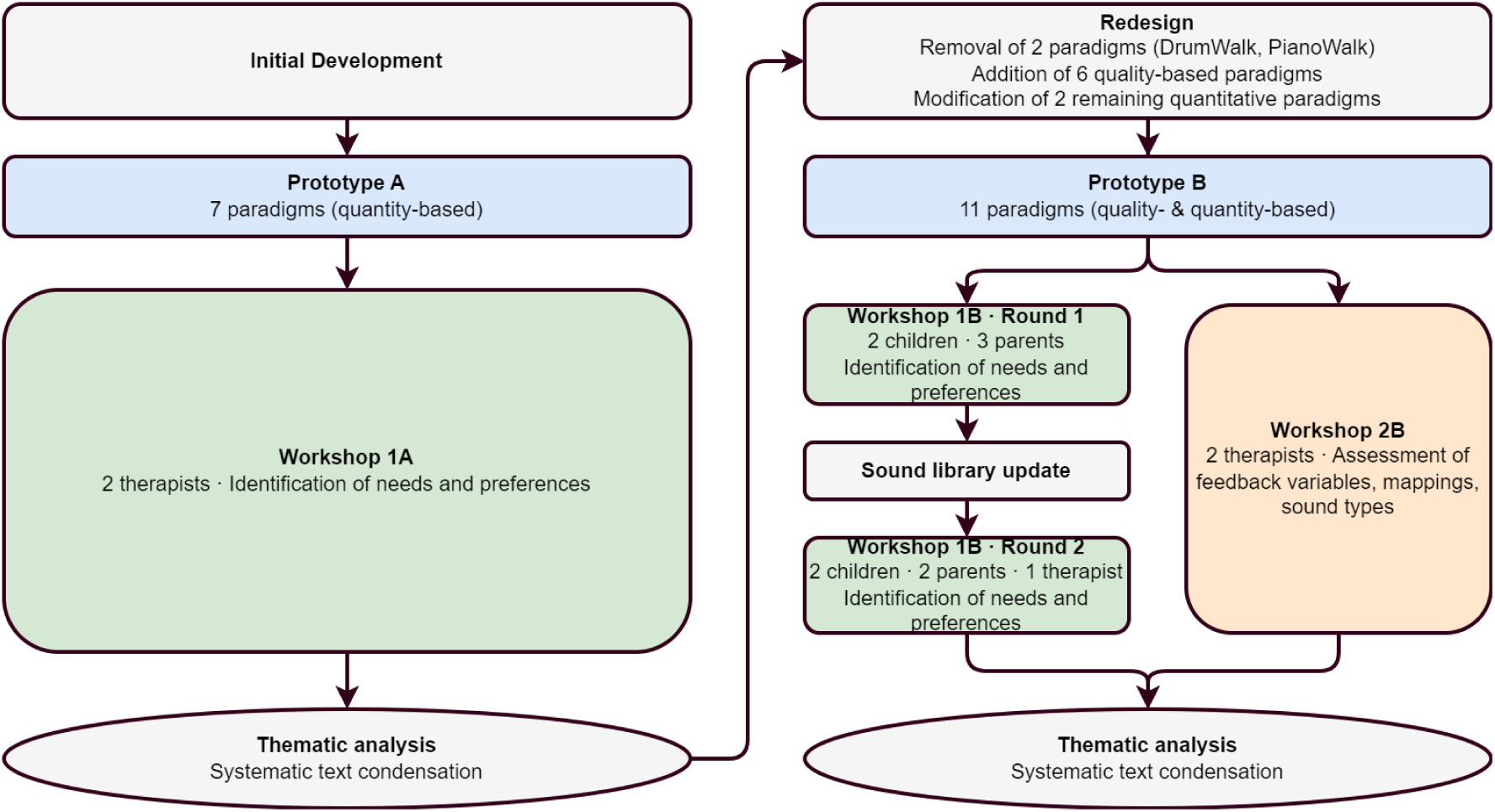
Activity flow across the two prototype phases.

### 2.2 Technology Description

The technological system that we used to develop the feedback paradigms was comprised of movement sensing hardware interfaced with a real-time software platform. The hardware component was composed of up to four wireless inertial measurement units (IMUs) (xIMU3, x-io Technologies, UK) that were placed on the lower extremities (foot, shin) using elastic velcro straps. Each unit was equipped with a gyroscope, a three-axis accelerometer, and a magnetometer, enabling registration of rotational velocity, linear acceleration, and orientation relative to the earth’s reference frame. The accelerometer and gyroscope data were integrated via x-io’s inbuilt sensor fusion algorithm, which calculated mediolateral and anteroposterior orientation with sufficiently high precision [47]. Acceleration, angular velocity, and orientation data were transmitted wirelessly via UDP protocol with a sampling frequency of approximately 160 Hz to a laptop computer. Transmission occurred through Wi-Fi using a high-performance router (xIMU Wireless Kit, 5 GHz, UK) to minimize latency, increase communication distance, and maximize data integrity. The feedback was played through a speaker (IKEA Eneby Gen 2, Sweden), connected to the computer via wired connection to ensure stable sound output with minimal latency.

Sensor data were received from the xIMU3 units by Mix-N-Map [48], a JUCE^1^-built software platform that interfaces with the sensors via the x-io Technologies C++ SDK^2^, reading IMU data at an internal callback frequency of 100 Hz. The platform is structured around the architecture of a multichannel audio mixer, in which each incoming data variable occupies a dedicated channel and can be subjected to a sequence of signal processing operations including threshold crossing detection, filtering, quantization, and nonlinear transfer function shaping. Processed channels can be normalized and combined before being routed to one of several audio synthesis engines: playback of pre-recorded sound samples, Markov chain-based algorithmic music generation, or real-time audio processing effects applied to existing audio streams. The mapping topology is fully configurable, supporting from one-to-one to many-to-many relationships between movement variables and sound parameters, with all mapping and processing parameters adjustable in real time during a session. This architecture lends itself directly to rapid prototyping of auditory feedback algorithms: a researcher or clinician can modify threshold values, transfer function shape, or the choice of audio engine on the fly, enabling iterative refinement of feedback paradigms within a single session and straightforward individualization to a given participant’s functional level and training needs.

### 2.3 Participants and Recruitment

Recruitment strategies differed by workshop type. For Workshop 1, children and their parents were recruited via a post in the Facebook group *CP Denmark*. The post described the study purpose, eligibility criteria (6-14 years with documented cerebral palsy, GMFCS levels 1-3), and contact information for interested parties. Therapists were recruited from two pediatric therapy clinics (1A municipal, 1B private) in Denmark. For Workshop 2, therapists were recruited from Elsass Fonden^3^. The participants across workshops are summarized below:

- Workshop 1A: Therapists 1 and 2, both female and with extensive CP-specific experience (12 and 28 years respectively), holding additional specialist certifications in pediatric neurological rehabilitation.
- Workshop 1B was conducted in two rounds. The first round included Child 1 (male, GMFCS level III, aged 5–9) and Child 2 (male, GMFCS level I, aged 5–9), accompanied by three parents. The second round included Child 3 (female, GMFCS level I, aged 5–9) and Child 4 (male, GMFCS level I, aged 10–14), each with one parent, and was also attended by an experienced physiotherapist (Therapist 5, female, 22 years of clinical experience).
- Workshop 2B: Therapist 3, a female physiotherapist with 10 years of CP-specific experience and a specialist certification in pediatric physiotherapy, and Therapist 4, a male occupational therapist with 4 years of CP-specific experience and broader neurological expertise from hospital and specialist pediatric settings.

### 2.4 Data Collection Procedures

All workshops were conducted in Danish and followed semi-structured formats designed to elicit participant perspectives on sound preferences and feedback paradigm utility. The specific procedures differed between Workshop 1 and 2, and also between therapist participants and child-parent participants within Workshop 1. Each workshop was staffed by members of the research team in four defined roles: a session lead responsible for receiving participants, introducing the workshop, and conducting consent procedures; an interview lead responsible for introducing each feedback paradigm, demonstrating the technology through walking and exercises, conducting the interview, and asking follow-up questions; a software operator responsible for managing the Mix-N-Map software during demonstrations and implementing real-time parameter adjustments in response to participant feedback; and a data recorder responsible for audio recording, timing, field notes, and secure data storage.

#### 2.4.1 Workshop 1A Procedure (Therapists, Prototype A)

Workshop 1A was designed to address RQ1 by eliciting sound preferences and design requirements from clinicians with direct experience of CP gait training, with a particular focus on which sound properties and feedback behaviors they judged appropriate and motivating for children with CP. The session lasted 90 minutes, with 15 minutes for project introduction and written informed consent, followed by approximately 60 minutes of paradigm demonstration and discussion at roughly 7 minutes per paradigm. The seven Prototype A paradigms (described in a subsequent section) were presented in a fixed order. For each paradigm, the interview lead wearing IMU sensors on the lower legs walked and performed CP-relevant gait training exercises, such as regular walking and stepping over obstacles. Therapists were invited to suggest specific movement variations during demonstrations. When participants proposed modifications, the software operator adjusted feedback parameters in real time wherever technically feasible, allowing therapists to evaluate proposed changes immediately rather than hypothetically. Discussion was guided by prepared questions displayed on slides when relevant topics were not raised spontaneously:

1. “How do you imagine that children might react to this sound?”
2. “Is there anything about this sound that you think could be particularly motivating or challenging for the children?”
3. “Is there anything about the sound that you think could be adjusted to better suit children with CP?”

#### 2.4.2 Workshop 1B Procedure (Children and parents, therapist, Prototype B)

Workshop 1B sessions were designed to address RQ1 by exploring which sound properties and feedback behaviors children with CP, their parents, and the involved therapists found engaging, motivating, or suitable for repeated gait training use. To minimize child fatigue, the sessions did not exceed 60 minutes, with 15 minutes allocated to informed consent procedures and familiarization. Approximately 30–35 minutes were then allocated to paradigm exploration, allowing roughly 5–6 minutes per paradigm. The demonstration procedure followed the same format as Workshop 1A, with the interview lead wearing IMU sensors and walking through exercises, but discussion questions were adapted for child participants:

1. “What do you think about the sound?”
2. “What do you like most about the sound?”
3. “ What do you not like about the sound?”
4. “Does it make you want to move?”
5. “Do the sounds make you feel happy, excited, nervous, or something else?”

During the session, the child was additionally encouraged to propose their own ideas for suitable or appealing sounds. Parents were invited to contribute observations on which sounds they thought would sustain their child’s motivation across repeated training sessions. Following the paradigm exploration phase, an optional second phase was conducted if the child had sufficient energy and was willing to participate. IMU sensors were placed on the child’s feet and lower legs, and the child walked back and forth in the room for up to one minute without auditory feedback, while the data recorder captured video and IMU data for use by the engineering team in future algorithm development. These data were not analyzed in the present study.

#### 2.4.3 Workshop 2B Procedure (Therapists, Prototype B)

Workshop 2B was designed to address RQ2 by having experienced physiotherapists assess which feedback variables, sound mapping structures, and sound types produced clinically viable and movement-interpretable auditory biofeedback for gait training across different GMFCS levels. The session lasted up to two hours, with 15 minutes for consent procedures and introduction followed by approximately 90 minutes of paradigm demonstration and discussion at roughly 15 minutes per paradigm. For each paradigm, a standardized introduction explained which movement parameter was measured and how sound behavior mapped to movement. The interview lead then demonstrated the paradigm by walking and performing gait training exercises with IMU sensors placed on the feet or lower legs, with sensor placement varying by paradigm. Following this, one therapist was invited to wear the IMU sensors and experience each paradigm firsthand through walking. Discussion was then facilitated using prepared questions on slides:

1. “How do you assess the paradigm in relation to its use in a therapeutic context?”
2. “How intuitive do you find the paradigm - both for you as a therapist and for the child with CP?”
3. “How do you perceive the pleasantness of the sound and its ability to convey relevant information in a therapeutic context?”

### 2.5 Data Analysis

All workshops were audio recorded and transcribed verbatim. Thematic analysis followed systematic text condensation [49]. The analysis process involved multiple readings of transcripts to achieve familiarization with the data, followed by systematic coding to identify meaning units. The codes were condensed into thematic categories, which in turn were synthesized into a narrative representation of the related findings. Two researchers independently coded the transcripts, with regular discussions to reach consensus on theme categorization and ensure internal consistency. Analysis continued until thematic saturation was achieved, indicating that no new themes emerged from the data. The final step was to translate the theme summaries and illustrative quotes to English with the help of Claude Sonnet 4.5^4^, which was followed by a manual sanity check.

### 2.6 Ethical Considerations

The project followed principles outlined in the Declaration of Helsinki and obtained approval from the North Denmark Region Committee on Health Research Ethics (ID: N-20240034). All participants received pertinent information about the study at least a day in advance and provided written informed consent prior to workshop participation. For child participants, consent was obtained from parents or legal guardians. All data were anonymized and stored securely according to Aalborg University’s GDPR-related policies.

## 3 Prototype A

Prototype A comprised a total of seven feedback paradigms, all of which were designed to encourage the maximization of the ‘amount of bodily movement per unit time’. This basic design choice aimed to support movement quantity-focused training approaches such as high-intensity interval training that are known to confer both functional and strength-related benefits in the CP population [50, 51]. The paradigms were realized through tailored combinations of movement-audio mappings that led to very distinct auditory experiences, but shared the following core aspects:

1. A normalized feedback variable, i.e. short-term movement quantity metric continuously computed in real time from shank-mounted inertial sensors.
2. Multivariate mappings of the feedback variable to sound generation / processing parameters that directly relate to the notion of ‘sonic intensity’, such as playback rate, music tempo, musical event density, loudness, and spectral brightness among others.

The movement quantity metric was developed and iteratively honed in the visual interface of Mix-N-Map until it was deemed to serve as a faithful approximation of short-term movement intensity variations during gait.

### 3.1 Quantity-Focused Feedback on Shank Rotation - Signal Processing Chain

Angular orientation of the shank segment in the anterior-posterior direction was received bilaterally from inertial measurement units (IMUs) mounted to the left and right shanks. The raw angular orientation signal from each limb, with a physical range of ±135 *^◦^*, was linearly normalized to the unit interval [0, 1] prior to processing:

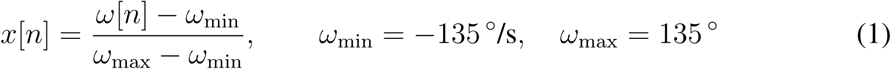

The normalized signal from each limb was passed through an identical, serially-chained processing pipeline. The first stage applied a second-order Butterworth low-pass filter implemented as cascaded biquadratic sections (two input delay elements, two feed-back delay elements each), cutoff frequency of *f_c,_* _LP_ = 7 Hz (*Q* = 0.7), operating at a sample rate of *f_s_* = 100 Hz. This configuration retained the frequency content relevant to walking kinematics [52]. The filtered signal was passed to a movement accumulator. At each sample *n*, the instantaneous movement increment was computed as the absolute first difference of the filtered signal *x̃*[*n*]:

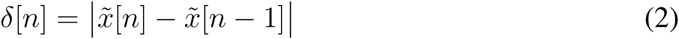

A rolling buffer retained the most recent *W* = 200 samples, corresponding to a 2-second window. The accumulated movement over the buffer was computed as a weighted sum with a quadratic gain ramp that progressively emphasized more recent samples:

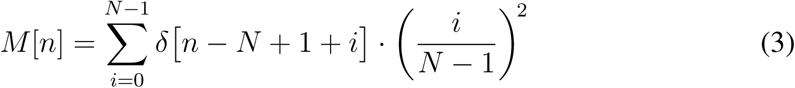

where *N* ≤ *W* denotes the current number of samples in the buffer. The accumulated value was then divided by a scalar normalization factor *α* = 5.0 and clipped to the unit interval, yielding the per-limb activity signal:

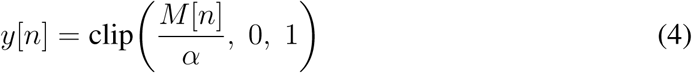

The processed outputs from both limbs, *y*_L_[*n*] (left) and *y*_R_[*n*] (right), were routed through a mapping matrix using an additive combination rule. Both channels contributed to a single intermediate output node with a gain factor *G* applied to the combined signal:

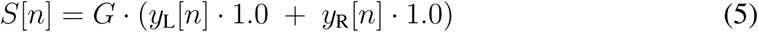

*G* could be directly adjusted in Mix-N-Map and controlled the sensitivity of the audio parameter changes to the activity signals. This intermediate node hence served exclusively as a bilateral activity aggregate, combining and scaling symmetric contributions from both limbs. The summed signal *S*[*n*] was subsequently distributed as the common input to one or more downstream output channels, each applying independent post-processing and parameter scaling for audio rendering.

### 3.2 Feedback Paradigms

Prototype A included a set of auditory feedback options, all of which involved concurrently mapping the aforementioned quantity metric to specific sound generation/processing parameters.

1. **FavoriteSong**: The interaction principle was to have the child control the playback speed of their favorite song such that the song only played back at its normal speed if the quantity metric continuously exceeded a set threshold, which could, for instance, be achieved by walking at a specific speed. Below the threshold, the song was slowed down, distorted, and bandlimited in the frequency domain to make the audible changes as salient as possible.
2. **StoryWalk**: This had the same interaction principle as FavoriteSong, with the only difference being that instead of a song, a story narration was used. The goal was to have the child ‘drive’ the narration forward through sustained movement. The narrations themselves could either be voice-only or with added light background music for mood-setting.
3. **VoiceWalk**: The principle here was to have the child control the articulatory properties of a funny synthesized human voice. The measured quantity metric controlled the synthesis parameters of the *Pink Trombone*^5^ vocal model (reimplemented as a module within Mix-N-Map) such that a greater quantity metric led to higher pitch and faster syllabic changes, and vice versa.
4. **MotorWalk**: The principle was to have a car motor sound that directly reflected the quantity metric. Smaller measurements led to a lower-pitched humming, which smoothly increased in pitch and intensity for progressively higher measurements.
5. **FartWalk**: The principle was that the child controlled a looped and pitch-modulated set of human flatulence sounds. When the child was stationary, there was no sound, and as the child exhibited more short-term movement, the fart sounds would become louder, higher in pitch, and faster.
6. **DrumWalk**: The principle was to have a generative drum performance whose moment-to-moment dynamics varied as a function of the quantity metric. This was achieved by mapping the latter to the parameters (event probability, velocity, and randomization) of a probabilistic drum synthesizer as implemented in [53]. The result was that smaller measurements led to a softer and less expressive performance, and larger measurements resulted in more drum hits per second and a greater level of musical intensity overall.
7. **PianoWalk**: The same principle as DrumWalk except that instead of drums, a generative piano model based on first-order Markov chains was used to create the musical output.

Demo video clips corresponding to the aforementioned paradigms are provided in an online folder^6^ with the exception of PianoWalk due to its interactional similarity to DrumWalk.

### 3.3 Results from Workshop 1A

Thematic analysis of Workshop 1A identified six themes: three relating to sound preferences and three relating to feedback paradigm design requirements. The former themes bear primarily on the sound library and how audio stimuli should be constructed to be motivating and comprehensible, while the latter address how feedback should be structured and delivered to support motor learning.

#### 3.3.1 Theme 1: Use of Recognizable Sounds

Therapists emphasized that employing familiar, recognizable sounds from children’s everyday environments could strengthen the connection between movement and auditory feedback, making training more meaningful and motivating. Recognizable sounds from contexts such as vehicles, animals, or popular media universes / franchises were perceived as more engaging than abstract or unfamiliar sounds.

The therapists explained that when children heard sounds they already knew and had positive associations with, they were more likely to engage actively with the training activity. One therapist specifically suggested that character voices from popular children’s media could be particularly motivating:

> *Can you find a sound that resembles this one [… ], from a known character from a cartoon?*

This principle extended to narrative content. The same therapist observed that children’s attachment to familiar stories could be harnessed directly:

> *If you have a favourite story, many children listen to the same one again and again and again because they love that story.*

This emphasis on recognizability also extended to sounds with motor-relevant associations. The motor sounds in MotorWalk were seen as intuitive because children could understand the connection between their movement (generating speed and power) and the motor revving. However, therapists stressed that what constitutes a “recognizable” or motivating sound varies substantially between individual children, depending on their age, interests, and life experiences. The technology therefore needed to offer a diverse library of sound options, and ideally should allow personalization where children or families could select sounds aligned with the child’s specific interests.

#### 3.3.2 Theme 2: Simple and Clear Soundscape for Effective Feedback

Therapists identified that auditory feedback needed to maintain acoustic clarity and simplicity to effectively support motor learning. When sounds were excessively complex or time-varying by nature, the therapists explained that children might struggle to understand which specific aspect of their movement had caused which aspect of the sound, undermining the feedback’s utility. This theme emerged most prominently in response to DrumWalk, where the model-intrinsic musical variations in articulation and timbre was perceived as obscuring the movement-sound relationship. One therapist articulated this from the perspective of clinical instruction:

> *If I as a therapist were to get a child to do something specific, I think it would be harder. There are too many different sounds and such. It is too complicated for a non-musical brain like mine at least.*

The same concern surfaced in response to StoryWalk, where the background music around the story narration was identified as unnecessary:

> *There doesn’t need to be all that surrounding sound, I think. [… ] I think it can be kept even simpler.*

In response to FartWalk, in which continuous quantitative feedback produced dense, varied sound output, the therapist acknowledged the engaging character of the paradigm while identifying that clarity was being sacrificed:

> *I think it is super fun, but it just needs to be much less, so you get a clearer response.*

In contrast, paradigms like MotorWalk, where there was a clear two-state distinction (motor off versus motor running), were seen as providing unambiguous information that children could readily interpret and use to guide movement adjustments. Therapists suggested that while some acoustic variation could be motivating, the core sound-movement mapping needed to remain simple and consistent enough that children could easily perceive cause-and-effect relationships between their actions and the auditory consequences.

#### 3.3.3 Theme 3: Avoid Monotony Through Sound Variation and Reward Mechanisms

While emphasizing the importance of simplicity and clarity (Theme 2), therapists simultaneously recognized that using identical sounds continuously throughout a training session would likely lead to habituation, boredom, and reduced engagement. Despite their generative nature, DrumWalk and PianoWalk both prompted explicit concern about this. Regarding DrumWalk, one therapist expressed worry about whether the presence of drums or piano alone could sustain a child’s attention:

> *It gets a bit too monotonous. I can be a bit worried [… ] will it become too monotonous compared to a favourite song?*

> *If you were to imagine putting something like this on, sensors, and perform treadmill-based gait training over time, this would quickly become boring.*

Conversely, conditions where the sensitivity of the sound mapping could be reduced so that the child’s movement produced more varied output were perceived as more engaging. As one therapist noted after experimenting with VoiceWalk at a lower configured sensitivity:

> *[… ] so it needs to go far down in sensitivity — then it becomes more fun. It was much more fun at the end.*

Therapists proposed that variation could also be achieved by offering multiple sound options within the same paradigm structure, allowing therapists or children to change sounds between exercises or sessions. Additionally, they suggested implementing reward mechanisms where achieving specific milestones triggered special sounds:

> *Every time you had walked for 30 seconds, you would get a reward for it, and then there’d be a little honk. Or whatever. Just something like that.*

This type of intermittent reinforcement through surprising or celebratory reward sounds was seen as potentially powerful for maintaining motivation, particularly for children who find repetitive motor practice tedious. The challenge identified was balancing this desire for variation with the need for clarity from Theme 2, ensuring that the basic movement-sound relationship remained understandable even when incorporating additional motivational elements.

#### 3.3.4 Theme 4: Auditory Feedback as Support for Movement Quality

In addition to sound preferences, workshop discussion also substantively addressed how feedback should be structured, from which three further themes were identified. The first of these concerned the distinction between quality and quantity as feedback targets. Therapists discussed auditory feedback as a means of ensuring qualitative correctness of movement, not merely quantitative performance. Heel strike was identified as the central gait parameter in therapeutic practice with this population:

> *Heel strike is the headline for these children. That is at least what we work a lot with, as therapists.*

When shown a video of an earlier version of FartWalk using an event-based qualitative paradigm in which the sound was contingent on heel contact rather than overall movement quantity, both therapists responded with clear approval. The exchange captured their reaction:

> *Therapist 2: That is really good.*
>
> *Therapist 1: And then it says nothing when the heel doesn’t come down?*
>
> *Software operator: No — if you walk on your toes, there is silence.*
>
> *Therapist 1: Yes, that is really good.*
>
> *Therapist 2: We would like to have that.*

This response indicated the need for integrating both quantitative and qualitative paradigms, as therapists recognized the value of quality-focused approaches while also noting how quantitative paradigms can help drive intensity. Having both types of paradigms available makes it possible to address different therapeutic goals in training for children with CP, supporting both movement quality and endurance/aerobic capacity.

#### 3.3.5 Theme 5: Adjustable Thresholds for Individualized Feedback

Therapists emphasized the practical importance of being able to adjust threshold and boundary values within the system. Without this capability, feedback would inevitably not accommodate variability in walking ability among slower or less able children with CP, meaning the positive feedback signal would be permanently inaccessible to them. One therapist described the problem explicitly with reference to FavoriteSong:

> *I think it is at least important to be able to know [… ] that even if it is perhaps a slow walker who would never be able to walk that fast [… ] you can turn down the threshold [… ] Otherwise they will never get up and get the good sound if they have to match our pace.*

This requirement applied both to lower and upper boundaries. Therapists saw adjustable limits as essential not only for matching the child’s current functional level but also for supporting goal-directed training, where thresholds could be progressively raised as gait function improved. Without such flexibility, the technology would risk being motivating only for the higher-functioning end of the target population.

#### 3.3.6 Theme 6: Immediate Sound Response for a Sense of Coherence

Therapists stressed that the temporal precision of the feedback response was a prerequisite for meaningful use. Delayed or lagging sound output would disrupt the perceived connection between movement and its auditory consequence, which they judged particularly problematic for children with cognitive challenges who might struggle to bridge even short temporal gaps between action and feedback. In response to DrumWalk, one therapist stated:

> *I don’t think it works very well. If it is to work, there needs to be at least a quick response when you slow down and speed up your walking pace.*

The importance of minimizing latency was also raised in relation to VoiceWalk:

> *And if some delay can also be removed — if that is even possible [… ] — then you get a somewhat faster and more enjoyable response.*

Therapists did not consider this purely a technical constraint to be accepted, but rather a design priority to be actively addressed in subsequent iterations.

## 4 Prototype B

### 4.1 Modifications Implemented in Prototype B

A core modification to the prototype based on the results from Workshop 1A (particularly Theme 4) was the addition of quality-focused feedback paradigms that specifically targeted heel strike. Prototype B hence comprised a combination of quality- and quantity-based feedback paradigms. The foundation of the quality-based approach was a signal processing chain that took as input the anteroposterior orientation of both feet and supplied as output a set of control signals for sound mapping. The key interaction principle was that the child needed to dorsiflex their ankles to cross a configurable angular threshold in order for the respective sound to be audible. The resulting feedback paradigms could either be:

1. Discrete, where crossing the minimum threshold triggered (typically) impulsive / short-duration sounds in a fixed or random sequence, or
2. Continuous, where a more steady state sound was amplitude modulated by the normalized foot angle, such that the minimum dorsiflexion threshold corresponded to the threshold of audibility, and angles exceeding the threshold led to sounds whose duration and intensity mirrored the duration and extent of dorsiflexion.

The signal processing chain applied to the raw foot orientation data is presented next.

#### Quality-Focused Feedback on Foot Orientation - Signal Processing Chain

The continuous and discrete paradigms both sourced bilateral estimates of foot segment orientation in the anterior-posterior plane from foot-mounted IMUs. The raw orientation signal from each limb, spanning a physical range of ±135*^◦^*, was linearly normalized prior to processing. The normalizer was configured to map a functional sub-range of the sensor output (dictated by the child’s range of motion) to [0, 1], discarding extreme values outside typical gait kinematics. The normalized signal from each limb passed through an identical two-stage processing pipeline, shared across both paradigms.

##### Stage 1: Low-pass Filtering

A second-order Butterworth low-pass filter was applied, implemented as cascaded biquad sections with a low-pass cutoff of *f_c,_* _LP_ = 7 Hz (*Q* = 0.7, *f_s_* = 100 Hz). This removed rapid incidental variations and high-frequency noise, retaining the kinematic content associated with the foot trajectory during gait [52].

##### Stage 2: Threshold Detection

The filtered signal *x̃_k_*[*n*] (*k* ∈ {L, R}) was passed through a threshold operation that computed the signed exceedance above a configurable upper bound *θ_H_* :

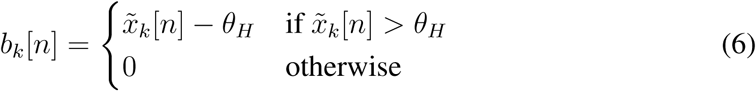

The output *b_k_*[*n*] was therefore a continuous-valued signal reflecting how far the foot orientation exceeded the threshold at each sample, and zero when it did not. The two paradigms drew on this signal differently: the discrete paradigm treated any nonzero value as a binary trigger event, discarding the magnitude, while the continuous paradigm used the magnitude directly to modulate audio parameters in proportion to the degree of dorsi-flexion above threshold.

##### Bilateral Summation

The per-limb signals resulting from the previous stage were combined with equal weighting via additive summation:

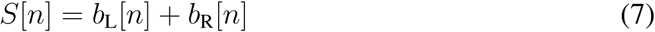

This bilateral aggregation treated the two limbs equally, and given the alternating nature of gait, only one limb ended up generating a nonzero signal value at any given point in time in practice.

##### Continuous Mapping

In the continuous paradigm, *S*[*n*] was passed through an asymmetric envelope follower with zero attack time and a finite release time constant (*τ_r_* ≈ 400 ms):

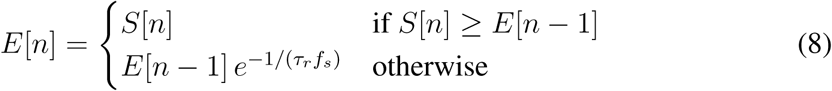

The envelope followed threshold exceedance instantaneously on the rising edge but decayed exponentially during inactivity, converting the sparse bilateral event stream into a smooth continuous signal that reflected recent gait activity. This signal continuously modulated the volume (gain) applied to a steady-state sound, such that the degree of foot orientation activity produced a graded, perceptually smooth effect.

##### Discrete Event Mapping

In the discrete paradigm, *S*[*n*] was routed directly to a trigger-type audio parameter, so that each moment at which at least one foot exceeded the threshold immediately fired a time-locked sound event. The sound level was held fixed and independent of the movement signal, and variation was achieved through the use of a sound folder with multiple short clips that could be triggered either serially or randomly.

#### 4.1.1 Overview of Feedback Paradigm Changes

From the original paradigm set, StoryWalk was retained without modification. Several paradigms were removed or refined, and five new quality-focused ones were added as described below (some before Round 1 of Workshop 1B, some after - see Fig. 1).

##### Removal

1. **DrumWalk** and **PianoWalk**: Both paradigms were removed entirely. The variable pitch and complex musical qualities produced acoustically rich but informationally ambiguous feedback (Theme 2), undermining the feedback’s utility for motor learning.

##### New Quality-Focused Paradigms

1. **CountWalk**: A discrete ‘step-counting’ paradigm that had a male voice counting up (1 - 2 - 3 - 4 - 1 - 2 - 3 - 4….) each time the user achieved sufficient ankle dorsiflexion during gait.
2. **AnimatedWalk TNT**: This was a variant of CountWalk with discrete Minecraft-inspired sounds triggered instead, with the goal of rewarding children using sounds that they were familiar with and positively disposed towards (added after Round 1 of workshop 1B).
3. **SafariWalk**: A third discrete variant of CountWalk which simulated a safari-type walking tour with discrete footsteps on rough terrain triggered by successful steps and a random animal vocalization triggered every 10th successful step (added after Round 1 of workshop 1B)
4. **FartWalk V2**: FartWalk was modified to function as a continuous quality-focused paradigm, where the same continuously looped fart sounds were modulated by the continuous foot angle trajectory as mentioned previously.
5. **CreakWalk**: Similar to FartWalk V2 making use of a continuous creaking.
6. **Cheering**: Also similar to FartWalk V2 making use of a continuous crowd cheering sound.

##### Refinement of Quantity-Based Paradigms

A general refinement applied to the previous paradigms was the adjustment of the rolling buffer window size from 200 samples to 100 samples to reduce the perceived latency of the sound changes in response to movement.

1. **FavoriteSong**: Expanded from a single user-selected song to a library of multiple options, preserving the simple threshold-based structure valued under Theme 2 while addressing the monotony concern of Theme 3 through greater variety of sound choices.
2. **MotorWalk**: The acoustic contrast between the sonic responses to smaller and larger movement quantity measurements was sharpened using two separate sounds to produce a more unambiguous two-state distinction, directly addressing the Theme 2 requirement for a perceptually clear movement-sound relationship.

Demo videos illustrating the new and revised paradigms in Prototype B are similarly provided in the same online folder with the exceptions of:

1. FavoriteSong: as the only change was related to expansion of the audio file library.
2. VoiceWalk: as the only change was related to the timbral parameters of the voice model and not the interaction.

### 4.2 Results from Workshops 1B and 2B

Prototype B was used across three workshop sessions. Workshop 1B was conducted in two separate rounds, in individual sessions per child, while Workshop 2B was run once. Thematic analysis across all sessions yielded nine themes: five derived primarily from Workshop 1B concerning sound preferences and paradigm proposals among children and parents, and four from Workshop 2B concerning paradigm evaluation by the therapists.

#### 4.2.1 Theme 1: Recognizable Sounds and Familiar Contexts as Motivational Anchors

Across both rounds of Workshop 1B, children responded most readily to sounds drawn from worlds they already inhabited, whether from video games, cartoons, or everyday physical environments. When a sound carried clear associations with a familiar context, they engaged with the paradigm immediately and without prompting, whereas acoustically unfamiliar or contextually unanchored sounds produced puzzled or lukewarm reactions. MotorWalk prompted a spontaneous response, captured in an exchange with Child 1:

> *Child 1: [… ] maybe for now, it’s a bit cool. Interviewer: Yes, a bit cool, OK. Parent 1: Do you think it’s cool? Child 1: Yes, I think maybe a little, yes.*

In the first round, both children also engaged actively when Minecraft-inspired sounds were discussed as potential library additions, and Child 1 responded with immediate enthusiasm to FartWalk. The second round amplified and specified these observations. During the demonstration, Child 3 spontaneously adopted an analogy the interviewer had mentioned earlier, reframing SafariWalk within a Pokémon-style game world. She described how the interaction could unfold:

> *If you walk around, you can pretend you’re in a Pokémon world, and then you walk into Pikachu, and it says Pikachu.*

Child 3’s mother explained that a particular suggestion had already come up at home before the workshop:

> *Child 3: Minions. Interviewer: Minions! Parent: When they say ‘Pa Poy!’ or ‘Ta Daa, Banana.’*

The pattern across both rounds was consistent: sounds with ready-made cultural associations triggered immediate affective responses, while VoiceWalk, whose synthesized voice lacked a clear referent, drew comparatively cautious or confused reactions from children.

#### 4.2.2 Theme 2: Individual Customization and Sensory Tolerance Boundaries

While familiarity was broadly motivating, what counted as familiar or pleasant varied substantially between the individual children. Preferences diverged not only in terms of which sound contexts were appealing but also in terms of which sounds fell within each child’s sensory tolerance, with the latter placing hard limits on what could function as positive reinforcement regardless of its general appeal. In the second round of workshops, certain sounds were rejected outright, including repeated dog barking and a counting voice, with participants describing them as irritating or intrusive rather than merely un-interesting. Customization requirements also intersected with therapeutic goals in ways that varied between families. Parent 1 identified a recurring problem in which the most motivating sound for her child was also the one most likely to drive counterproductive movement:

> *[…] a bit of the problem with you, child, that when you train, you end up being a motorcycle. And often you need to be a turtle… You like to run and be fast, but there’s a lot of focus on getting the movements done correctly.*

Therapist 4 addressed customization from a clinical perspective when evaluating Count-Walk, framing individualization (in the form of calibrating the target range of motion) not as a convenience feature but as the central design principle:

> *I think the more you can customize to each individual, and that’s both in relation to the range of motion we talked about before, but also in relation to the sounds, so that you go into their world and say OK, there’s one here who loves Pokémon. Then try to find all those sounds from Pokémon.*

Taken together, the findings from both workshops point to customization as a layered requirement: the sound library must be broad enough to accommodate diverse interests, the sensory profile of each option must stay within individual tolerance, and the character of the sound must be compatible with the movement characteristics the therapist is trying to reinforce.

#### 4.2.3 Theme 3: Situational and State-Dependent Adaptation

Data from the second round of Workshop 1B introduced a temporal dimension to the customization requirement that was not clearly visible in the first round: sound preferences within individual children were not fixed but shifted with energy level and momentary state. A paradigm that worked well for a child on an energetic day could be unsuitable after a demanding school afternoon, meaning that even a well-calibrated individual sound profile would require revision from session to session. MotorWalk prompted an immediate energizing response from Child 3 on the day of her workshop:

> *Physiotherapist 1: So we can pretend to be a racecar down the hall, right? Child: Then I’d really hurry up! Parent: Yes, then you’d hurry — run fast!*

Yet the same parent reflected that this would not always be the right choice. On days following a tiring school day, the energy-driving quality of MotorWalk could become a liability rather than an asset, and the FavoriteSong paradigm was proposed as an alternative precisely because of its different character:

> *Sometimes, if you’re a little tired when you have to train, it can sometimes help to have a bit of music.*

The practical consequence is that switching between paradigms during or between sessions must be fast and low-effort enough to happen responsively, not only as a result of advance planning by the therapist.

#### 4.2.4 Theme 4: Library Expansion, Sound Differentiation, and Competition Elements

Children and parents in both rounds proposed extensions to the sound library and modifications to existing paradigms oriented toward greater specificity and a higher ceiling for sustained engagement. The proposals converged on a common concern: that a library limited in scope or consisting of sounds without clear contextual grounding would exhaust a child’s interest relatively quickly, and that certain paradigms set the bar for reward too low to motivate effort across a full session.

VoiceWalk attracted particular criticism on the grounds of acoustic strangeness. Child 1 described its synthesized voice quality as “a bit jöøh,” a rendering of something neither recognizable nor classifiable. In contrast, Child 2 responded positively when MotorWalk was discussed in terms of a specific sound he associated with a game he already played:

> *Software operator: But would it make sense for it to sound like a tractor or a motorcycle, maybe? Child 2: A tractor. Parent 3: Yes, you like that one. Child 2: It’s the same in Farming Simulator.*

Regarding CountWalk, participants in both rounds noted that the step count ceiling (four) was too low to sustain engagement across a session. Children proposed extending the count to 100 or even 1000 steps, reflecting an intuition that reaching a larger goal carries more reward than reaching a small one quickly. The suggestion of milestone-triggered explosion or ignition sounds from the AnimatedWalk TNT sounds at specific step counts, raised by Child 2, expressed the same logic and received support from the workshop group:

> *There I’d rather have a creeper that blows up. Or some TNT that you can just hear ignite.*

#### 4.2.5 Theme 5: Alternative Paradigms for Controlled Tempo and Inverted Feedback

Both rounds of Workshop 1B produced spontaneous proposals for paradigm structures not present in Prototype B, all involving sound contingent on restraint or controlled movement rather than on speed or effort. These suggestions emerged without prompting and reflected a perception that the existing paradigm set rewarded exertion predominantly, leaving therapeutic goals involving slow, deliberate motor execution without a corresponding feedback structure. Parent 1 proposed an inversion of FartWalk: rather than a sound triggered by heel strike, the paradigm would play a continuous sound by default and silence it only when the child moved quietly and carefully, framing the task as a game of controlled, deliberate footfall:

> *And it could actually be quite fun if it was reversed. So that there was sound and then you had to be quiet, and you could only be quiet if you crept.*

FavoriteSong prompted a closely related proposal, this time inverting the threshold relationship of the speed-based playback paradigm. When the interview lead suggested that the song could play at its best quality during slow walking and degrade as pace exceeded a target, the exchange captured both the child’s immediate comprehension and the parent’s endorsement:

> *Interview lead: Whether you could reverse it, so it plays well if you walk slowly, and if you walk too fast then it doesn’t sound very good. Could that be something you think could be fun to walk with? Child 1: I think so. I think so. Parent 1: Actually, I think you’d find it fun to play with tempo like that and get to manipulate the sound.*

Both proposals point toward a category of feedback paradigm in which the therapeutic target is modulation downward rather than upward, with potential relevance for gait training goals involving controlled speed, deliberate heel contact, or reduced co-activation.

#### 4.2.6 Theme 6: Flexible Feedback for Children’s Unpredictable and Creative Responses

Therapists in Workshop 2B emphasized that children with CP frequently approach feed-back systems through routes outside any anticipated use pattern, and that this unpredictability should be treated as a property to design for rather than a deviation to eliminate. When a child uses a novel movement strategy to activate a feedback paradigm, that invention can itself represent productive therapeutic engagement, provided the system is open enough to reward it. Therapist 4 illustrated this with reference to FartWalk, describing the range of unexpected strategies a child might develop to trigger the heel strike feedback:

> *How children react to it… There I just have to keep coming back to this thing about them having a completely different frame of understanding than I do as an adult. There may be some who lie down and try to do something with their feet in the air, or some who start crawling on the wall [ladder]… Sort of trying to figure out their thoughts on how do I activate this sound, and what does it do for me?*

Therapist 3 framed this orientation explicitly as a clinical value:

> *I usually say we love when children outsmart us — if they find a different solution than sitting and rocking their foot.*

The design implication is that paradigms should not be constructed so narrowly that only one movement strategy can activate the feedback. An open-ended structure allows children to engage on their own terms initially, with the therapist progressively guiding them toward the target movement pattern as the session advances.

#### 4.2.7 Theme 7: Flow and Rhythm as Integrating Principles

Therapists identified a distinct experiential quality in certain paradigms that could be described as flow: a sense of continuous, meaningful integration between movement and sound in which the activity felt self-sustaining rather than requiring repeated cognitive effort to maintain. This quality was attributed primarily to FavoriteSong, where the coupling of walking pace to musical playback speed created a feedback loop (‘flow’) that Therapist 4 found more compelling in practice than paradigms providing explicit, movement-specific auditory cues:

> *It makes more sense to me with the flow than with the specific, concrete feedback on a movement. The flow feedback makes more sense to me.*

Beyond individual training, therapists recognized that this property opened possibilities for embedding the paradigm within group-based exercises. Therapist 4 described a catch game in which the speed of the shared music could serve as a signal regulating the pace of all participants simultaneously:

> *[… ] You could make a game of catch where when the sound plays slowly, everyone has to walk more slowly, and run faster or something. There are many things you could embed this in.*

Rhythmic, flow-based paradigms thus appear to have affordances that extend beyond individual biofeedback toward group motor rehabilitation contexts, potentially increasing both the clinical flexibility and the social engagement value of the technology.

#### 4.2.8 Theme 8: Intuitive Sound-Movement Connection

Therapists evaluated paradigms partly on whether the relationship between movement and sound would be immediately self-evident to a child encountering the system for the first time. Where this connection could be grasped without verbal explanation, therapists judged the paradigm as having strong clinical utility: less time would be required to orient each child at the start of a session, and the cognitive resources freed by an intuitive mapping could be directed toward the motor task itself.

FartWalk, redesigned in Prototype B to trigger sound at discrete heel strike events rather than varying continuously with movement speed, was identified by both therapists as achieving this quality. Therapist 4’s assessment following its demonstration was direct:

> *I think it works very intuitively, for me. This thing about it being clear feed-back that I do something, then something happens, and then I get a sound I recognize. Yes. In that way I think it’s fine.*

Therapist 3 confirmed the same judgment from the child’s perspective: “For the child, it’s quite intuitive.” The underlying criterion was the reliability and simplicity of the cause-effect relationship: a child who could form a clear mental model of which specific action produced which specific sound was equipped to use that understanding to guide intentional movement adjustments.

#### 4.2.9 Theme 9: Functional Contexts and Sense of Mastery

Therapists argued that feedback paradigms are most effective clinically when connected to activities that give children a concrete, bodily sense of what they are capable of, rather than framing training as the correction of a movement deficit. The distinction carries practical implications both for how paradigms are presented to children and for which movement parameters are chosen as feedback targets. Therapist 3 developed this point in relation to CountWalk, drawing on her assessment of how many children with CP relate to training for isolated movement components:

> *I probably wouldn’t find myself saying I’m doing intensive toe-walking training with a child, but I could find it relevant for them to understand what their body can do, and there you’d have something like having to hit something with the foot or the heel, walk uphill, so you combine it in a game or something, so that they get the connection that they can actually activate the muscles, and for very many children it is also about them being tight in the back of the legs and it being hard to activate the front. One thing is that it’s tight and that’s just how it is. But one could be curious about how much they can activate.*

The argument is that embedding joint-level feedback within activities that resemble something the child already does or could aspire to do in daily life preserves the therapeutic specificity of the feedback while giving the movement intrinsic meaning. Walking uphill, navigating an obstacle course, or playing a game involving targeted foot placement can each carry the same ankle feedback that would otherwise feel like isolated remedial exercise, but within a context that generates a sense of mastery rather than deficit.

### 4.3 Summary of Recommended Modifications

Based on these themes, several design priorities for further development were identified across both workshop types. The sound library should be substantially expanded, with Pokémon and Minion character vocalizations specifically requested by Children 3 and 4, and milestone-triggered reward sequences added to SafariWalk at fixed step counts. CountWalk’s step ceiling should be extended to accommodate longer training bouts with accumulating goals. FavoriteSong should be modified so that volume decreases in parallel with playback speed when movement drops below threshold, to accommodate children with sound sensitivity. Inverse-threshold paradigms, where sound rewards slow and controlled movement rather than speed, should be prototyped in direct response to the demand observed across both rounds of Workshop 1B. The interface should support rapid in-session switching between paradigms to enable day-to-day adaptation to the child’s state.

## 5 Discussion

This study set out to identify, in the context of auditory biofeedback technology, the needs and sound preferences of children with CP, and to establish which structural properties must be fulfilled by feedback paradigms in order to support gait training in this population. The findings across the two prototype iterations helped define a set of design constraints grounded in the specific clinical and developmental characteristics of CP: which feedback variables are therapeutically appropriate, which mapping structures remain interpretable under those constraints, and which sound types sustain engagement without obscuring the movement-sound relationship. The ensuing discussion interprets each cluster of findings in that light.

### 5.1 Within-Child Preference Variability and Its Implications for System Architecture

The finding that sound preferences fluctuated within individual children depending on energy level and sensory state reinforces the notion that user profiling at the individual level is not a sufficient basis for personalization. Many past biofeedback systems had been designed around stable user characteristics (i.e., a child would be assessed, a configuration was then selected, and that configuration was applied during the training session and possibly across sessions as well) [20]. The present data suggest that this model is inadequate for children with CP, whose sensory processing difficulties [15, 54] and variable attentional capacity [24, 42] make within-child state variation a clinically relevant phenomenon rather than minor temporal variability around a stable preference. Session-level adaptability must be a core design requirement, not a ‘nice-to-have’ feature. This also has consequences for how future effectiveness trials should be designed. If engagement quality fluctuates with the child’s state, aggregate outcome metrics averaged across sessions will obscure the relationship between feedback appropriateness and motor learning. Structured session-level monitoring of engagement is methodologically necessary if the contribution of auditory feedback to training outcomes is to be isolated.

### 5.2 Sound-Movement Coherence as a Functional Constraint on Feedback Complexity

The consistent preference for paradigms with interpretable movement-sound relationships, and the corresponding rejection of the generative musical paradigms in Prototype A, can be understood in terms of the basic requirements of augmented feedback for motor learning. For feedback to drive movement correction, the learner must be able to attribute the feedback signal to a specific property of their movement [31]. This attribution requires that the mapping between action and auditory consequence be both perceptible and unambiguous. In children with CP, sensorimotor integration difficulties [18, 54] already compromise the internal feedback signals that typically guide movement, placing a higher burden on the clarity of the external feedback signal. The therapists’ responses to DrumWalk and PianoWalk in Workshop 1A suggest that generative musical paradigms violate this requirement not because they are acoustically complex per se, but because their complexity is not tied to movement in a way the child can decode. FartWalk in its Prototype B form demonstrates the complementary point: a culturally anchored, acoustically distinct sound can be both engaging and fully interpretable when it maps discretely to a single gait event. The design constraint is therefore not acoustic simplicity as such, but what might be called informational transparency: the child must be able to form a reliable mental model of which aspect of their movement produces which change in sound. This reframes the tension between engaging sound design and effective feedback as a tractable engineering problem.

### 5.3 Inverted Feedback, Structural Bias of the Existing Paradigms

The spontaneous and unprompted requests for feedback paradigms rewarding restrained and controlled movement, emerging independently across Workshop 1B parents, point to something more significant than idiosyncratic preference. They reveal a structural bias in the existing auditory biofeedback literature for CP: the documented paradigm space is almost exclusively organized around increasing movement output, whether in terms of step frequency, speed, or amplitude [20, 31, 33]. This orientation reflects the historic emphasis on hypokinetic gait deviations in the biofeedback literature, but it does not cover the full range of therapeutic targets encountered in clinical practice. Children whose primary goals involve controlled deceleration, larger range of motion at a slow pace, or modulated co-activation require a feedback structure in which doing less, or doing it more carefully, produces the reward. The inverse paradigm proposals in Workshop 1B define this requirement precisely at the design level. These proposals underscore the value of including family participants in co-design processes since they have direct experience of their child’s training program, and identifies inverse-threshold paradigms as a concrete and clinically justified priority for the next development phase.

### 5.4 Motoric Creativity, Rhythmic Flow, and Functional Framing

Three further themes from Workshop 2B address properties of feedback paradigms that bear on how children with CP engage with training over time, and connect to established principles in CP rehabilitation. The therapist observation that children’s unanticipated motor strategies for activating a feedback system constitute productive engagement rather than protocol deviation is consistent with the broader emphasis in motor rehabilitation on movement variability as a resource rather than an error [55]. A paradigm that can only be activated by one precisely specified motor pattern will suppress this variability; a paradigm built around a clear but open-ended trigger allows the therapist to shape motor exploration progressively toward the clinical target. This has direct implications for how feedback paradigms are specified during design: over-constraining the trigger can hence become a barrier to engagement. The flow property identified in FavoriteSong, in which continuous coupling between walking pace and musical playback created a self-sustaining feedback loop, describes a different but complementary mechanism. Where event-based paradigms provide discrete information about specific movement events, flow-based paradigms may sustain the training intensity and duration necessary for motor adaptation without requiring the child to maintain explicit attentional focus on a feedback signal. These two paradigm types are not in competition; the therapist preference for a repertoire spanning both quality- and quantity-based feedback reflects a clinically accurate recognition that different training goals require different feedback structures. Finally, the argument for embedding joint-level feedback within functional activities is reinforced by (1) the consistency with which joint kinematic adaptations are seen in CP-specific biofeedback literature [30], and (2) the therapist observation that the concrete framing of the training (see Theme 9 - Functional Contexts and Sense of Mastery) affects the child’s relationship to the activity. Whether this has measurable effects on training adherence or long-term outcomes is an empirical question that the present data cannot answer, but it provides a theoretically coherent rationale for task-embedded feedback paradigm design.

### 5.5 Comparison with Existing Literature

The positive participant responses to clearly structured event-based paradigms are broadly consistent with Baram and Lenger’s demonstration that simple auditory feedback achieves effects on gait parameters comparable to visual biofeedback [33]. The present data extend that finding by showing that simple click sounds [33] represent one design point in a wider space of potential feedback designs that is naturally constrained by cultural anchoring, feedback polarity, and sound-movement coherence. Schoenmaker et al.’s observation [31] that effect sizes in Hamed and Abdelwahab’s music-based auditory feedback study [34] exceeded those of Baram and Lenger’s click-based approach [33] supports the potential of the present findings, with the caveat that this comparison is cross-study rather than controlled. The present findings suggest that musical richness and feedback effectiveness are not straightforwardly related: participants responded positively to FavoriteSong and rejected DrumWalk and PianoWalk, not on acoustic complexity per se, but on whether the sound remained interpretable as movement-contingent feedback. The relevant dimension is hence not acoustic simplicity or complexity in themselves, but whether the child can interpret the feedback as informative about a specific aspect of their movement. More generally, a paradigm can be acoustically varied and culturally rich while remaining interpretable, as the FartWalk evaluation in Workshop 2B demonstrated; it can also be acoustically sophisticated while being informationally ambiguous, as DrumWalk and PianoWalk illustrated in Prototype A.

The iterative workshop approach operationalizes the user-centered development recommendations of Graham et al. [2] and Wheeler et al. [39] in a domain where end-user involvement has been consistently called for but is rarely documented in detail. The tensions that the process brought to light, particularly between short-term feedback predictability and novelty, illustrate why neither user testing alone nor clinician specification alone is sufficient to guide design. The FartWalk redesign from speed-contingent to heel-strike-contingent feedback illustrates how early therapist input can shape subsequent child-facing evaluation, though the non-overlapping therapist cohorts across the two evaluation rounds mean that direct comparisons of therapist responses between prototypes should be treated with caution. Our finding regarding the dependence of feedback preferences on the mental / physical state of the child at the time of training, which was not previously reported [31], [20], [30], suggest that future effectiveness trials should include structured monitoring of engagement quality across sessions.

### 5.6 Limitations and Future Directions

The convenience sample was small and functionally restricted: all four child participants were at GMFCS levels I or III, leaving children with more severe motor impairments, younger children, and adolescents entirely unrepresented. The design principles identified here should therefore be understood as grounded hypotheses for a specific functional range rather than general requirements for the full CP population, and their generalizability will need to be assessed in studies with a broader population. A further limitation is lack of data saturation in both rounds of workshops with therapists; however, the therapists involved had ample experience working with children with CP: Workshop 1A was conducted with therapists holding 12 and 28 years of CP-specific experience, while Workshop 1B was conducted with a single therapist with 22 years of experience and Workshop 2B involved therapists with 10 and 4 years respectively. Workshops were conducted in a controlled clinical setting rather than during actual training sessions, and short-term novelty effects at first exposure to a technology (such as with virtual reality [56]) are difficult to separate from genuine preference. Whether the preferences identified here would sustain engagement over weeks of repeated daily use is unknown and must be investigated through longitudinal feasibility studies before effectiveness trials can be adequately designed. Generally speaking, the workshops involved small convenience samples of therapists ranging in experience, and the results should therefore be interpreted cautiously.

No objective movement data were analyzed, and participant preferences cannot therefore be interpreted as indicators of motor learning efficacy. This is an inherent constraint of the design phase rather than a methodological shortcoming [43]: it would have been premature to evaluate gait outcomes for paradigms that had not yet been validated for basic usability. The appropriate next step is a within-subject comparison of participant-preferred paradigms in a structured training context, measuring both gait parameters and engagement quality. Simplified sensor attachment solutions and a caretaker-friendly interface supporting in-session paradigm switching and adjustment are development priorities that must be addressed before larger-scale clinical or home-based evaluation can begin.

## 6 Conclusions

Through the direct involvement of relevant stakeholders, this user-centered design study identified key requirements and preferences for auditory biofeedback technology for gait training among children with CP. This work not only yielded insight into which sound types are preferred by children in the Danish context, but also showed that preferences appear to vary within individual children depending on their energy level and sensory state on a given day. This makes the child-session combination, rather than the individual alone, a key factor dictating feedback algorithm choice and subsequent individualization, which directly influences system architectural decisions. Specifically, it points to the need for session-level interface capabilities to support individualized mappings, movement range calibration and rapid modification of the sonic palette for biofeedback generation.

Several more specific conclusions follow from the thematic analysis. Sound-movement coherence functions as a binding constraint on feedback complexity: the necessary removal of the layered generative musical paradigms demonstrated that acoustic richness becomes counterproductive once it obscures the interpretability of the movement-sound relationship. Qualitative paradigms targeting specific movement events (e.g. heel strike) were consistently preferred by therapists on clinical grounds and are well-justified by the sensorimotor integration deficits that make augmented feedback relevant for this population. The spontaneous demand from participants for ‘inverted’ paradigms rewarding restrained and controlled movement identifies a feedback category that does not yet exist in the current prototype and has received little attention in the auditory biofeedback literature for CP more broadly.

The iterative user-centered process generated findings that may have been missed by a specification-driven development approach. The inverted paradigm proposals, the therapist responses to heel-strike-contingent feedback, and the clinical reframing of children’s unpredictable motor strategies as therapeutically valuable all emerged through direct participant engagement with the working technology. This encourages the continued involvement of children, families, and therapists in subsequent development phases. Future research should combine objective gait outcome measurement with structured monitoring of engagement quality across sessions, as the present data suggest that engagement fluctuates with the child’s state in ways that aggregate outcome metrics alone will not capture. This is crucial as engagement is a key mediator of training adherence, which in turn directly dictates long-term movement outcomes and quality of life in children with CP.

## Data Availability

All data produced in the present study are available upon reasonable request to the authors

## Data Availability Statement

The transcription and coding data underlying the findings of this study are not publicly available owing to the privacy and confidentiality protections afforded to participants under the approved research protocol.

## Acknowledgments

The authors gratefully acknowledge the children, parents, and therapists who participated in this study. This work was supported by the Elsass Foundation under Grant 23-B01-1400.

## Disclosure of AI use

Large language model-based AI tools were used during the preparation of this manuscript to support initial drafting (translating parts of the originally Danish-written report to English, optimizing textual framing of some introductory arguments) and to revise specific passages for clarity and conciseness. AI was also used for partial automatic transcription of the therapists’ interviews; all transcripts were manually reviewed and corrected by the authors. AI was further used as language and methodological support during the systematic text condensation process, while all analytic decisions remained the responsibility of the researchers. All content was critically reviewed and edited by the authors, who take full responsibility for the accuracy and integrity of the published work.

## Footnotes

1 https://www.juce.com

2 https://github.com/xioTechnologies/x-IMU3-Software/tree/main/Examples/Cpp

3 https://www.elsassfonden.dk/

4 https://www.anthropic.com/news/claude-sonnet-4-5

5 https://github.com/imaginary/pink-trombone/

6 https://drive.google.com/drive/folders/18T_fUpH56Vi9OzICCIDtMQQjdlyMxfsB

## References

[1] P. Rosenbaum, N. Paneth, A. Leviton, M. Goldstein, M. Bax, D. Damiano, et al. A report: the definition and classification of cerebral palsy april 2006. Developmental Medicine & Child Neurology Supplement, 109:8–14, 2007.

[2] H. K. Graham, P. Rosenbaum, N. Paneth, B. Dan, J. P. Lin, D. L. Damiano, et al. Cerebral palsy. Nature Reviews Disease Primers, 2:15082, 2016.

[3] M. Oskoui, F. Coutinho, J. Dykeman, N. Jetté, and T. Pringsheim. An update on the prevalence of cerebral palsy: a systematic review and meta-analysis. Developmental Medicine & Child Neurology, 55(6):509–519, 2013.

[4] M. Pirpiris, P. E. Gates, J. J. McCarthy, J. D’Astous, C. Tylkowksi, J. O. Sanders, et al. Function and well-being in ambulatory children with cerebral palsy. Journal of Pediatric Orthopaedics, 26(1):119–126, 2006.

[5] Gaelle E. Doucet, Sarah Baker, Tony W. Wilson, and Max J. Kurz. Weaker connectivity of the cortical networks is linked with the uncharacteristic gait in youth with cerebral palsy. Brain Sciences, 11(8):1065, 2021.

[6] Yinglu Hong, Dapeng Bao, Brad Manor, and Junhong Zhou. Characterizing the supraspinal sensorimotor control of walking using MRI-compatible system: a systematic review. Journal of NeuroEngineering and Rehabilitation, 21(1):34, 2024.

[7] Alain Frigon, Turgay Akay, and Boris I. Prilutsky. Control of mammalian locomotion by somatosensory feedback. Comprehensive Physiology, 12(1):2877–2947, 2022.

[8] Frédéric J. F. Viseux. The sensory role of the sole of the foot: Review and update on clinical perspectives. Neurophysiologie Clinique, 50(1):55–68, 2020.

[9] Serge Rossignol, Réjean Dubuc, and Jean-Pierre Gossard. Dynamic sensorimotor interactions in locomotion. Physiological reviews, 86(1):89–154, 2006.

[10] R. Frost, J. Skidmore, M. Santello, and P. Artemiadis. Sensorimotor control of gait: a novel approach for the study of the interplay of visual and proprioceptive feedback. Frontiers in Human Neuroscience, 9:14, 2015.

[11] C. D. MacKinnon. Sensorimotor anatomy of gait, balance, and falls. Handbook of Clinical Neurology, 159:3–26, 2018.

[12] Kaoru Takakusaki. Functional neuroanatomy for posture and gait control. Journal of Movement Disorders, 10(1):1–17, 2017.

[13] Michael P. Trevarrow, Anna Reelfs, Sarah E. Baker, Rashelle M. Hoffman, Tony W. Wilson, and Max J. Kurz. Spinal cord microstructural changes are connected with the aberrant sensorimotor cortical oscillatory activity in adults with cerebral palsy. Scientific Reports, 12:4807, 2022.

[14] Germana Cappellini, Yury P. Ivanenko, Giovanni Martino, Michael J. MacLellan, Annalisa Sacco, Daniela Morelli, and Francesco Lacquaniti. Immature spinal locomotor output in children with cerebral palsy. Frontiers in Physiology, 7:478, 2016.

[15] A. Mahesan, P. Jauhari, M. Singhal, S. Gulati, B. Chakrabarty, V. Sirolia, et al. A relook at cerebral palsy beyond motor pathology: A cross-sectional study of sensory processing abilities. Neurology India, 72(3):590–596, 2024.

[16] M. K. A. Barakat, G. H. Elmeniawy, and F. H. Abdelazeim. Sensory systems processing in children with spastic cerebral palsy: a pilot study. Bulletin of Faculty of Physical Therapy, 26(1):27, 2021.

[17] ME Busse, Charles Mark Wiles, and Robert William Martin Van Deursen. Muscle co-activation in neurological conditions. Physical Therapy Reviews, 10(4):247–253, 2005.

[18] J. Lorentzen, M. Willerslev-Olsen, H. Hüche Larsen, S. F. Farmer, and J. B. Nielsen. Maturation of feedforward toe walking motor program is impaired in children with cerebral palsy. Brain, 142(3):526–541, 2019.

[19] S. Dhiman, S. Kundra, N. Jha, R. K. Goyal, P. Ajmera, and S. Gulati. Sensory processing issues in children with cerebral palsy: A systematic review. Current Developmental Disorders Reports, 12(1):9–, 2025.

[20] A. MacIntosh, E. Lam, V. Vigneron, N. Vignais, and E. Biddiss. Biofeedback interventions for individuals with cerebral palsy: a systematic review. Disability and Rehabilitation, 41(20):2369–2391, 2019.

[21] A. T. C. Booth, A. I. Buizer, P. Meyns, I. L. B. Oude Lansink, F. Steenbrink, and M. M. van der Krogt. The efficacy of functional gait training in children and young adults with cerebral palsy: a systematic review and meta-analysis. Developmental Medicine & Child Neurology, 60(9):866–883, 2018.

[22] Marika Demers, Karen Fung, Sandeep K. Subramanian, Martin Lemay, and Maxime T. Robert. Integration of motor learning principles into virtual reality interventions for individuals with cerebral palsy: Systematic review. JMIR Serious Games, 9(2):e23822, 2021.

[23] N. G. Moreau, A. W. Bodkin, K. Bjornson, A. Hobbs, M. Soileau, and K. Lahasky. Effectiveness of rehabilitation interventions to improve gait speed in children with cerebral palsy: Systematic review and meta-analysis. Physical Therapy, 96(12):1938–1954, 2016.

[24] D. P. Moss. The use of biofeedback and neurofeedback in pediatric care. Functional Symptoms in Pediatric Disease: A Clinical Guide, pages 285–303, 2014.

[25] Lara Martínez-Rodríguez, Cristina García-Bravo, Sara García-Bravo, María Salcedo-Pérez-Juana, and Jorge Pérez-Corrales. New technological approaches in occupational therapy for pediatric cerebral palsy: A systematic review. In Healthcare, volume 13, page 459. MDPI, 2025.

[26] Roland Sigrist, Georg Rauter, Robert Riener, and Peter Wolf. Augmented visual, auditory, haptic, and multimodal feedback in motor learning: a review. Psychonomic Bulletin & Review, 20(1):21–53, 2013.

[27] A. M. Spomer, B. C. Conner, M. H. Schwartz, Z. F. Lerner, and K. M. Steele. Multisession adaptation to audiovisual and sensorimotor biofeedback is heterogeneous among adolescents with cerebral palsy. PLOS ONE, 19(11):e0313617, 2024.

[28] L. van Gelder, A. T. C. Booth, I. van de Port, A. I. Buizer, J. Harlaar, and M. M. van der Krogt. Real-time feedback to improve gait in children with cerebral palsy. Gait & Posture, 52:76–82, 2017.

[29] E. Dursun, N. Dursun, and D. Alican. Effects of biofeedback treatment on gait in children with cerebral palsy. Disability and Rehabilitation, 26(2):116–120, 2004.

[30] Leonie Hirsch and Natalie Mrachacz-Kersting. Augmented feedback as a therapeutic approach for gait rehabilitation in patients with cerebral palsy: A systematic review. Frontiers in Rehabilitation Sciences, 7:1638091, 2026.

[31] Jorine Schoenmaker, Han Houdijk, Bert Steenbergen, Heleen A ReindersMesselink, and Marina M Schoemaker. Effectiveness of different extrinsic feed-back forms on motor learning in children with cerebral palsy: a systematic review. Disability and Rehabilitation, 45(8):1271–1284, 2023.

[32] Antonia Zaferiou, Zahava Hirsch, Tristan Bacani, and Luke Dahl. A review of concurrent sonified biofeedback in balance and gait training. Journal of NeuroEngineering and Rehabilitation, 22(1):38, 2025.

[33] Y. Baram and R. Lenger. Gait improvement in patients with cerebral palsy by visual and auditory feedback. Neuromodulation, 15(1):48–52, 2012.

[34] N. S. Hamed and M. S. Abdelwahab. Pedometer-based gait training in children with spastic hemiparetic cerebral palsy: a randomized controlled study. Clinical Rehabilitation, 25(2):157–165, 2011.

[35] Tamara Veronica Pinos Cisneros, Annette Brons, Ben Kröse, Ben Schouten, and Geke Ludden. Playfulness and new technologies in hand therapy for children with cerebral palsy: scoping review. JMIR Serious Games, 11(1):e44904, 2023.

[36] Gillian King, Helene Schwellnus, Dianne Russell, and Nicolette Terblanche. Engaging children with cerebral palsy in interactive computer play-based motor therapies: theoretical perspectives. Disability and Rehabilitation, 43(1):133–147, 2021.

[37] Naomi Gefen, Barbara Mazer, Tal Krasovsky, and Patrice L Weiss. Novel rehabilitation technologies in pediatric rehabilitation: knowledge towards translation. Disability and Rehabilitation: Assistive Technology, 20(5):1209–1218, 2025.

[38] S. G. S. Shah and I. Robinson. Benefits of and barriers to involving users in medical device technology development and evaluation. International Journal of Technology Assessment in Health Care, 23(1):131–137, 2007.

[39] G. Wheeler, N. Mills, U. Ankeny, P. Howsley, C. Bartlett, H. Elphick, et al. Meaningful involvement of children and young people in health technology development. Journal of Medical Engineering & Technology, 46(6):462–471, 2022.

[40] A. Majnemer, K. Shikako-Thomas, N. Chokron, M. Law, M. Shevell, G. Chilingaryan, et al. Leisure activity preferences for 6- to 12-year-old children with cerebral palsy. Developmental Medicine & Child Neurology, 52(2):167–173, 2010.

[41] I. Novak, C. Morgan, M. Fahey, M. Finch-Edmondson, C. Galea, A. Hines, et al. State of the evidence traffic lights 2019: Systematic review of interventions for preventing and treating children with cerebral palsy. Current Neurology and Neuroscience Reports, 20(2):3, 2020.

[42] P. R. Kantan, S. Dahl, and E. G. Spaich. Adapting audio mixing principles and tools to parameter mapping sonification design. Proceedings of the 29th International Conference on Auditory Display (ICAD 2024), pages 148–155, 2024.

[43] N. Bleijenberg, J. M. de Man-van Ginkel, J. C. A. Trappenburg, R. G. A. Ettema, C. G. Sino, N. Heim, et al. Increasing value and reducing waste by optimizing the development of complex interventions: Enriching the development phase of the medical research council (mrc) framework. International Journal of Nursing Studies, 79:86–93, 2018.

[44] S. K. Tatla, K. Sauve, N. Virji-Babul, L. Holsti, C. Butler, and H. F. M. Van Der Loos. Evidence for outcomes of motivational rehabilitation interventions for children and adolescents with cerebral palsy: an American Academy for Cerebral Palsy and Developmental Medicine systematic review. Developmental Medicine & Child Neurology, 55(7):593–601, 2013.

[45] Prithvi Ravi Kantan, Sofia Dahl, Helle Rovsing Jørgensen, and Erika G Spaich. Making movement sonification usable in clinical gait rehabilitation: A user-centered study. In Proceedings of the 19th International Audio Mostly Conference: Explorations in Sonic Cultures, pages 43–60, 2024.

[46] Prithvi Ravi Kantan, Sofia Dahl, Helle Rovsing Jørgensen, Chetali Khadye, and Erika G Spaich. Designing ecological auditory feedback on lower limb kinematics for hemiparetic gait training. Sensors, 23(8):3964, 2023.

[47] Sebastian OH Madgwick, Andrew JL Harrison, and Ravi Vaidyanathan. Estimation of IMU and MARG orientation using a gradient descent algorithm. In 2011 IEEE International Conference on Rehabilitation Robotics, pages 1–7. Ieee, 2011.

[48] Prithvi Ravi Kantan, Sofia Dahl, and Erika G Spaich. Adapting audio mixing principles and tools to parameter mapping sonification design. In The 29th International Conference for Auditory Display (ICAD 2024) The 29th International Conference on Auditory Display (ICAD 2024), 2024.

[49] Kirsti Malterud. Systematic text condensation: a strategy for qualitative analysis. Scandinavian Journal of Public Health, 40(8):795–805, 2012.

[50] Reidun Lauglo, Torstein Vik, Torarin Lamvik, Dorthe Stensvold, Ane-Kristine Finbråten, and Trine Moholdt. High-intensity interval training to improve fitness in children with cerebral palsy. BMJ Open Sport & Exercise Medicine, 2(1), 2016.

[51] Christian Schranz, Annika Kruse, Teresa Belohlavek, Gerhardt Steinwender, Markus Tilp, Thomas Pieber, and Martin Svehlik. Does home-based progressive resistance or high-intensity circuit training improve strength, function, activity or participation in children with cerebral palsy? Archives of physical medicine and rehabilitation, 99(12):2457–2464, 2018.

[52] Cesare Angeloni, Patrick O Riley, and David E Krebs. Frequency content of whole body gait kinematic data. IEEE Transactions on Rehabilitation Engineering, 2(1):40–46, 1994.

[53] Prithvi Ravi Kantan. Probabilistic generative music as a data sonification medium. In 3rd Conference on Sonification of Health and Environmental Data (SoniHED 2025), pages 77–84. KTH Royal Institute of Technology, 2025.

[54] H. Piitulainen, M. Sukanen, T. Finni, and F. Cenni. Proprioceptive-perception threshold is impaired in cerebral palsy and is associated with worse balance performance. Gait & Posture, 106:S165–S166, 2023.

[55] Ashesh K Dhawale, Maurice A Smith, and Bence P Ö lveczky. The role of variability in motor learning. Annual Review of Neuroscience, 40:479–498, 2017.

[56] Ines Miguel-Alonso, David Checa, Henar Guillen-Sanz, and Andres Bustillo. Evaluation of the novelty effect in immersive virtual reality learning experiences. Virtual Reality, 28(1):27, 2024.

